# Automated Enhancement and Curation of Healthcare Data

**DOI:** 10.1101/2022.04.03.22273359

**Authors:** Melody Greer, Cilia Zayas, Sudeepa Bhattacharyya

## Abstract

Social and behavioral aspects of our lives significantly impact our health, yet minimal social determinants of health (SDOH) data elements are collected in the healthcare system. In this study we developed a repeatable SDOH enrichment and integration process to incorporate dynamically evolving SDOH domain concepts from consumers into clinical data. This process included SDOH mapping, linking compiled consumer data, data quality analysis and preprocessing, storage, and visualization. Our preliminary analysis shows commercial consumer data can be a viable source of SDOH factor at an individual-level for clinical data thus providing a path for clinicians to improve patient treatment and care.

## 1 Introduction

Socioeconomic and behavioral aspects of our lives significantly impact our health, yet minimal social determinants of health (SDOH) data elements are collected in the healthcare system. Information of this type is needed for quality healthcare research and patient care because it is associated with the full-spectrum of health outcomes from acute to chronic disorders. Studies indicate cancer (Alcaraz et al. 2020), cardiovascular disease (Tamura et al. 2019), dementia (Nicholas et al. 2021), mental health and substance-abuse disorders (Galea and Vlahov 2002, Alegría et al. 2018), viral infection (Greer et al. 2021)and sleep (Grandner and Fernandez 2021) are among a long list of health problems (Kivimäki et al. 2020) which are linked to social risk factors not frequently or consistently collected for patients. The combined effect of missing, inconsistent, or inaccurate data also leads to bias in machine learning, algorithms underlying clinical decision support, predictive analytics or other healthcare processes. (Obermeyer et al. 2019, Cottrell et al. 2020, Seker, Talburt, and Greer 2022) To avoid these problems as well as to gain rich insights from healthcare data we must be cognizant about diverse data collection, veracity and data-quality.

Electronic health records (EHR) are assembled from clinical, insurance and basic demographic information during the course of patient care. Although there is growing interest in including SDOH data into EHR, the capture and management mechanisms for this process are uncertain.(Ancker et al. 2018, Feldman, Davlyatov, and Hall 2020, Gold et al. 2018) Current possibilities include (1) paper-based or digitally collected social needs screening before a patient receives care, (2) during a visit with a clinician, or (3) publicly available data sets that provide social context. All of these methods have challenges. Screening data that is incorporated into the EHR appears fragmented to users, increases the staff workload plus adds a data entry step where paper is used. (Gold et al. 2018) And although clinicians felt SDOH information was valuable they are already pressed for time, and adding to their list of clerical tasks is often not practical or even possible. (Tong et al. 2018) These barriers are likely the reason for the findings of Fraze et al. that social needs screening for food, housing, utilities, transportation, and experience with interpersonal violence was present in 24.4% of hospitals, and 15.6% of physician practices. (Fraze et al. 2019) SDOH data documented within unstructured EHR fields during a visit needs further study and will require natural language processing tools to be integrated into healthcare practice. (Hatef et al. 2019) Publicly available data, whether in raw form (i.e. Census), or aggregated into measures (i.e. SDI, ADI) has been valuable in studying the relationships between socioeconomic and patient health status (Chamberlain et al. 2020, Johnson-Lawrence, Zajacova, and Sneed 2017, Tung et al. 2018). However, appending community-level data introduces issues with averaging. In their 2020 work on community-level and patient-level social risk data Cottrell et al. observed that community-level data misses some patients that patient-level data would not. (Cottrell et al. 2020) Further complicating the issue, there are no currently accepted standard SDOH data elements. (Cantor and Thorpe 2018) This means that even in cases where social risk data is consistently collected it cannot be easily shared with other healthcare providers during transfers or referrals.

Compiled consumer data can address these issues. Finance and marketing businesses have successfully used this data for over twenty years to find customers, understand their needs, and tailor financial products. Using these same strategies, healthcare researchers and providers can improve patient treatment and care. The consumer data includes individual-level SDOH data providing a holistic snapshot of an individual’s lifestyle. It includes amongst others, income, education, lifestyle variables, language spoken, household size, smoking status, life events, hobbies, shopping activity etc. that are not available in the insurance claims data or majority of EHR data. A support system, for complex assessments (i.e., risk assessments) and calculations, is needed using information that helps to arrive at conclusions regarding patient’s health risk and treatment. The development of an automated pipeline process for multisource healthcare data integration will provide this support. The presence of information integrated from multiple different streams of data will support tools for nurses, social workers, community health workers and patient navigators that supports decision making by providing the ability to consider multiple factors simultaneously for patients and clinicians. To begin the development of an automated pipeline process for multisource healthcare data integration we have conducted a pilot study integrating clinical and consumer information sources to evaluate multiple SDOH and clinical factors simultaneously.

## 2 Methods

Our goal was to prototype a repeatable clinical data enhancement process to incorporate compiled consumer data into SDOH domain concepts. This process included social risk factor mapping, linking compiled consumer data, data quality assessment, preprocessing, storage, and visualization. Consumer data does not currently line up one-to-one with factors identified as SDOH, so purchased data elements were mapped to concepts identified in social needs screening. Once the mapping was complete, the compiled consumer data elements were linked to the patient population and stored in a Microsoft SQL Server. The resulting data was then accessible using SQL queries, and the quality was evaluated for completeness, consistency, and timeliness or temporal alignment. Visualization tools allow researchers or clinical decision support developers to study the data before analysis or application. Combining a SQL database with an academic license for Tableau provided easy access for visualization, data analysis, and wrangling. As a proof-of-concept, we provided SAS JMP to clinicians and researchers to determine what aspects of these tools deliver the most value. SAS JMP can be easily connected directly to the database and provides many descriptive analysis and visualization methods. Open source tools, like R and Python, can also serve the same purpose. Throughout SDOH enhancement, a security and data privacy layer overlaid the entire process with security features included in each step to ensure data privacy. Figure 1 depicts a snapshot of the entire process.

**Figure 1:**
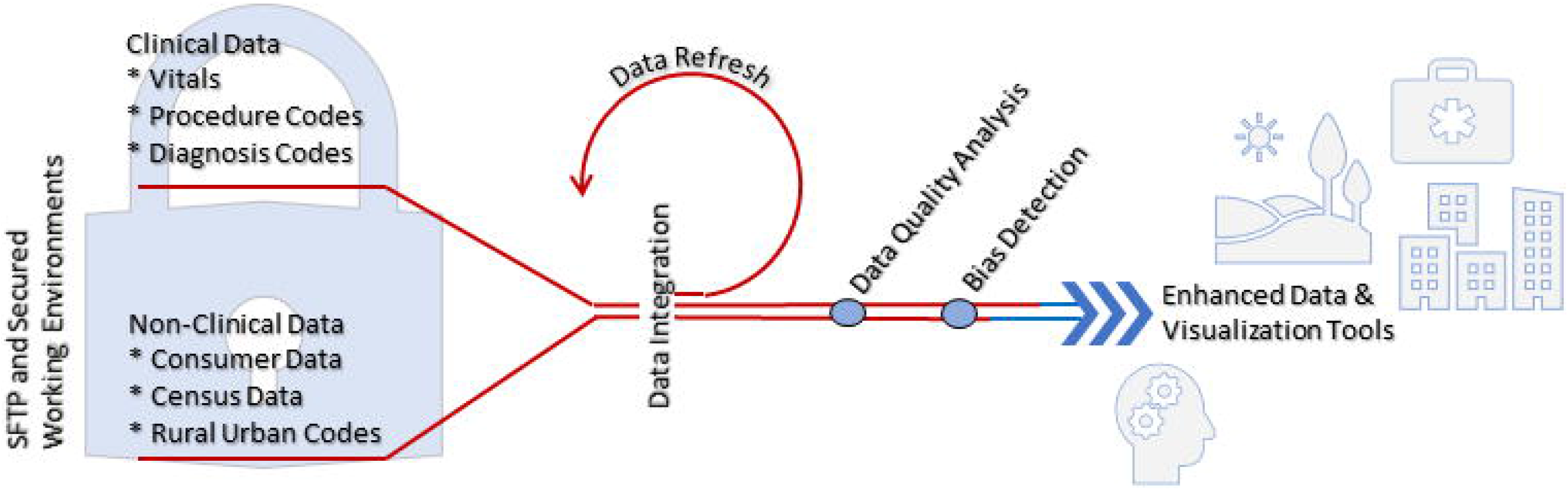
Workflow describing data integration of clinical and SDOH factors translated from consumer information sources.

### 2.1 Mapping

Various social determinants of health frameworks have been created to assist communities, healthcare professionals, and others in better identifying and managing an expansive range of factors influencing health outcomes. (Office of Disease Prevention and Health Promotion 2022a, World Health Organization 2010, Rural Health Information Hub 2022) Our mapping strategy was developed based on the Healthy People (HP) 2030 SDOH Framework for this study. (Office of Disease Prevention and Health Promotion 2022a, b) HP 2030 not only continued the HP initiative, which set national health targets for 2020 through 2030, but also designed a framework to organize SDOH into five domains: (1) economic stability, (2) education, (3) social and community context, (4) health and healthcare access, and (5) the neighborhood and built environment. HP 2030 outlined essential SDOH within each of these domains. (Office of Disease Prevention and Health Promotion 2022b) Employment, food insecurity, housing instability, and poverty, for example, all fall under the domain of economic stability. We mapped as many elements as possible based on the HP 2030 framework from the two consumer data sources.

### 2.2 Clinical Data

We requested electronic health records from the Clinical Data Repository (University of Arkansas for Medical Sciences Translational Research Institute 2022) for all patients with chronic conditions (i.e., asthma, diabetes, heart disease, congestive heart failure, coronary artery disease, heart attack, stroke), or contagious respiratory illness (i.e., influenza, or COVID-19). All data received was stored on one of the following secure devices: institute supported controlled access server, institute supported password protected desktop computer, encrypted password protected laptop. The data used for linking social determinants information was name, address, DOB only. These demographics were transmitted to the selected data compiler vendors via SFTP.As per the requirement of the Health Insurance Portability and Accountability Act of 1996 (HIPAA) for protection of patient health information these 3^rd^ party vendors signed a Business Associate Addendum (BAA) with our medical institution prior to accessing the patient identification. Following the addition of the SDoH the data was de-identified to increase protection of the participants from any negative consequences in the event of a data breach. De-identification was accomplished by deleting full name and address and replacing them with a random identification number and RUCCA code (cite). The DOB was deleted and replaced with age. Data was stored in a secure database server behind a firewall.

### 2.3 Non-Clinical Data Integration and Refresh Process

Commercial data is updated monthly and is made up of hundreds of different sources, including consumer surveys, public records, purchase transactions, real estate data, offline and online buying behavior, and warranty information. Wherever possible, compilers compare values from multiple data sources to check accuracy for each element. Vendors that compile data for commercial purposes (i.e., marketing) were identified and interviewed, and then costs were negotiated. SDoH data points were appended by commercial data compilers or other external data sources using only patient name, DOB, and address. These processes occur entirely within a database system using fully HIPAA compliant vendors. All data was encrypted while in transit and immediately destroyed at the compiler location after the completion of the processing. Not all clinical data will be matched to existing consumer data during this process. This is the problem of coverage which refers to the number of patients who could be linked to compiled data. Coverage varies by commercial compiler, but the reasons for coverage variation may also be associated with varying aspects of the patient’s lifestyle (e.g., people who use cash exclusively are less likely to have a substantial digital imprint in consumer databases).

### 2.4 Data Quality Analysis

Data quality is a constellation of factors essential for data collections. The quality of the linked clinical and commercial data (henceforth called merged-data) was evaluated for conformance, completeness, consistency, plausibility and temporal alignment. (Kahn et al. 2016) As was mentioned above, accuracy, and consistency, were evaluated by compilers before data was purchased. After data was purchased and linked with EHR data we further measured consistency of common data elements between compilers from the different sources. In the merged-data there were 5 data elements that were common between the compilers. We matched the data records based on these 5 data elements to measure consistency between the compilers as an added level of data-quality assessment. Each of the data elements differed in categories or levels, between the compilers. Our first step was to collapse the categories in each element into identical categories so that they could be compared for consistency. For example, the data element ‘Home Market Value, Estimated’ in compiler 1 mapped to ‘Home Value Range_VDS_Appended’ in compiler 2. The categories of each these feature variables were however not the same. ‘Home Market Value, Estimated’ in compiler 1 had the following 20 categories; ‘$1,000 - $24,999’, ‘$25,000 - $49,999’, ‘$100,000 - $124,999’, ‘$125,000 - $149,999’, ‘$150,000 - $174,999’, ‘$175,000 - $199,999’, ‘$200,000 - $224,999’, ‘$225,000 - $249,999’, ‘$250,000 - $274,999’, ‘$275,000 - $299,999’, ‘$300,000 - $349,999’, ‘$350,000 - $399,999’, ‘$400,000 - $449,999’, ‘$450,000 - $499,999’, ‘$50,000 - $74,999’, ‘$500,000 - $749,999’, ‘$75,000 - $99,999’, ‘$750,000 - $999,999’, ‘$1,000,000 Plus’, ‘NA’. The ‘Home Value Range_VDS_Appended’ in compiler 2 had the following 18 categories; ‘Under $50k’, ‘$50 - $100k’, ‘$100 - $150k’ ‘$150 - $200k’, ‘$200 - $250k’, ‘$250 - $300k’, ‘$300 - $350k’, ‘$350 - $400k’, ‘$400 - $450k’, ‘$450 - $500k’,’$500 - $550k’, ‘$550 - $600k’ ‘600 - $650k’,’$650 - $700k’, ‘NA’, ‘$700 - $750K’, ‘$750K +’, ‘Unknown’. In order to assess consistency we first collapsed the categories in each variable in an intuitive and meaningful fashion and made them identical. The new collapsed categories in both variables were: ‘Less100K’, ‘100-200K’, ‘200-300K’, ‘300-400K’, 400-500K, ‘500Kplus’. After discarding the ‘NA’s or ‘Unknowns’, the consistency between the two variables were calculated.

Missing data are a pervasive problem in any source of data also referred to as data ‘completeness’. Lack of complete data can significantly affect a study outcome by introducing unwanted bias. This is why it was important for us to obtain consumer marketing data from a diverse selection of sources to ensure that we have a collection of data that is complete and deep enough to provide meaningful information about majority of the study participants.

During preprocessing we also examined conformance and plausibility of the data elements, thus comparing the actual format of the data against the expected, and evaluating the feasibility of multiple existing values of the data elements. Measuring the persistence of the data was not possible in this work since to analyze changes in the data over time multiple batches of consumer data would be needed which was cost-prohibitive.

### 2.5 Bias

Human bias exists. As we collect, analyze and take actions based on data our biases are perpetuated. This bias pervades healthcare in machine learning, decision support, operations and logistics planning in health systems. Applications to guide clinical practice are not exempt. Real-world clinical data is important for clinical decision-making but it has everyday biases imprinted within it and can preserve or even amplify health disparities. To address this issue requires detection and correction. Sensitive attributes (i.e., race, gender, etc.) are evaluated against classifications to first detect bias that may be present. Once uncovered biased data can be rebalanced using class labels. While this method is not full proof, biases that are not expected may remain hidden, it does allow for the mitigation of known issues and for the discovery of unknown issues.

### 2.6 Data Analysis and Visualization

After the data was linked and preprocessed we explored the data using Tableau, SAS JMP and R statistical software packages that were linked to the Sql server hosting the merged-data. We determined the summary statistics of a subset of the variables, from both EHR and compiled SDOH sources, that characterized the patients. We are also currently in the process of building classifiers that can predict disease risk based on patients’ clinical, demographic and SDOH factors. There are open-source tools available such as R, Python and others, that can perform interactive data analysis and visualization at no cost and can be customized to the needs of clinicians. For example, Shiny is an R package that can be used to build dashboards or provide the clinicians or researchers a means to interact with the data as per their need. Our ongoing effort focuses on developing an R-Shiny based analysis and visualization tool as well.

## 3 Results

### 3.1 Mapping and Linking Consumer Data with EHR at an individual-level

**Table 1** lists SDOH domains, elements, and coverage percentages from two consumer data databases that were used in our study and named here as compiler 1 and compiler 2. There were 55,422 patients in the initial set of EHR records. After linking with two commercial compilers, 30,895 and 54,880 patients with SDOH data remained respectively. As the table shows, compiler 1 had fewer mapped data elements compared to compiler 2. However, compiler 1 had, on an average, ∼90% coverage on the mapped data elements while compiler 2 had many more elements mapped but the coverage was much lower, ∼54% on an average. Thus, consumer data collection from at least two sources ensured that all categories of SDOH domains, based on Healthy People 2020 SDOH Framework were covered in our merged-data. (Office of Disease Prevention and Health Promotion 2022a) The connection between unlinked patients’ needs further study to determine what they have in common aside from a minimal individual digital footprint.

**Table 1:**
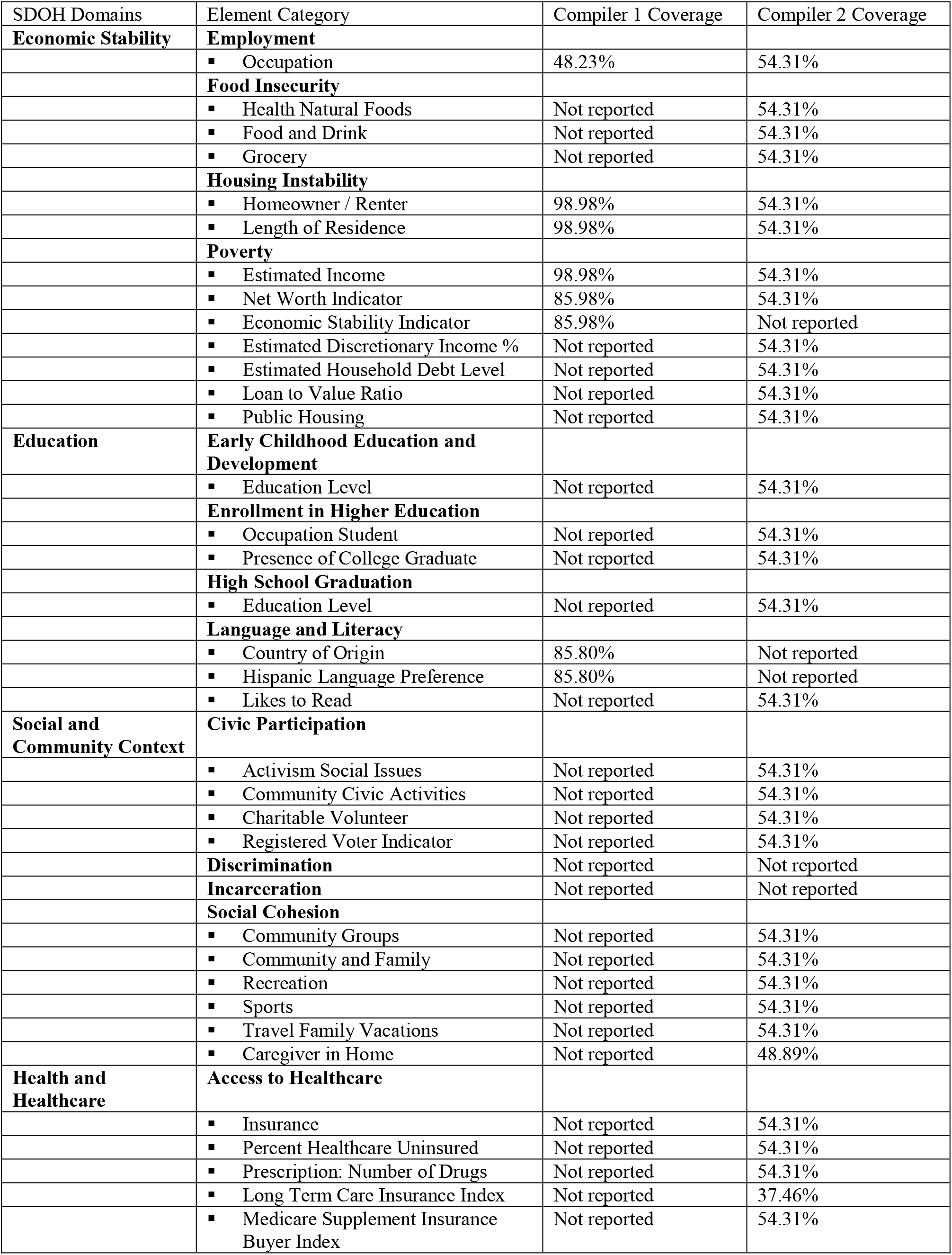
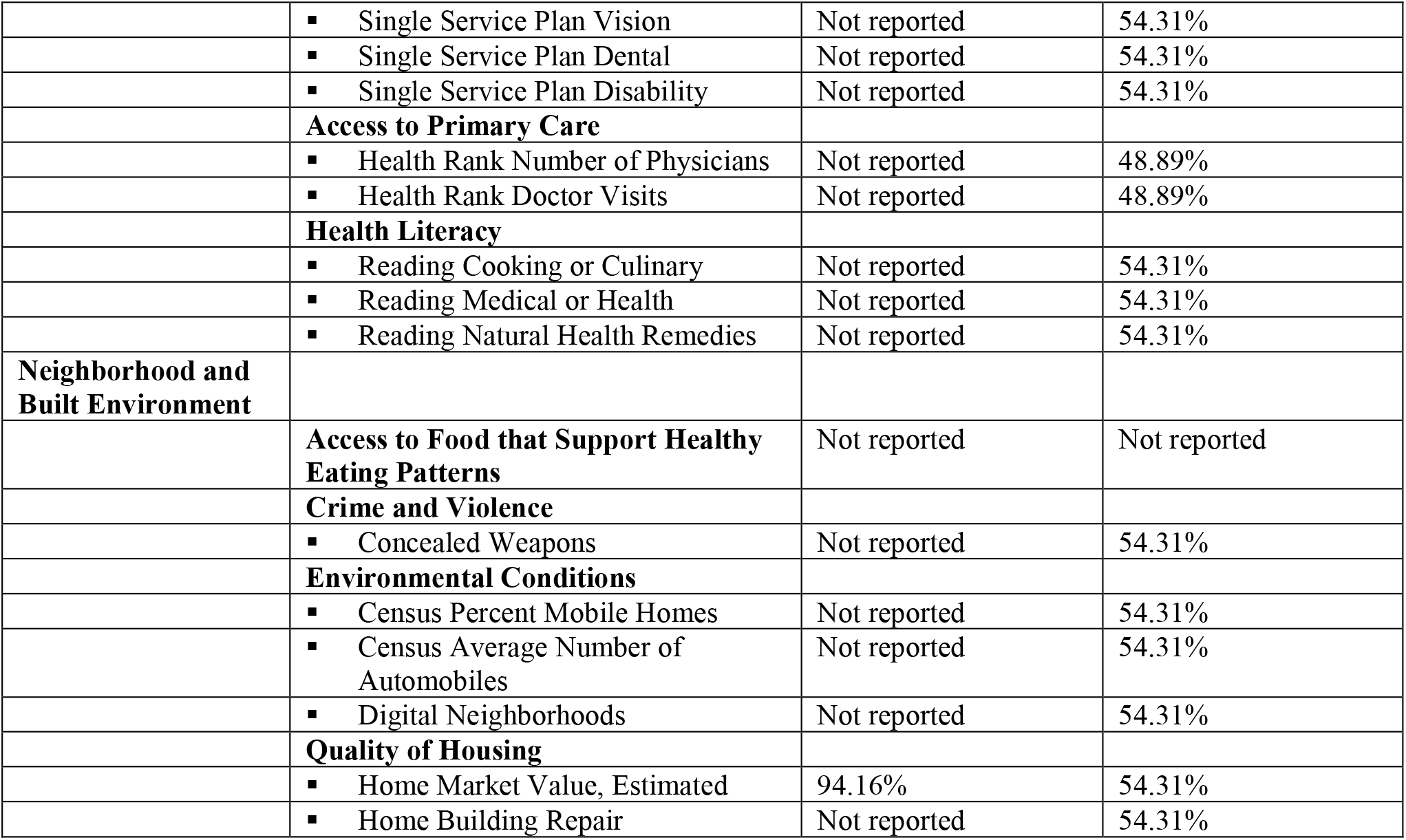
Data elements mapped from consumer databases to SDOH categories with coverage percentages listed for each individual compiler.

### 3.2 Data quality assessment and preprocessing

To measure consistency between the compiler data collected from different sources we matched the data records based on elements that were common between the compilers as part of data-quality assessment. **Table 2** shows the randomly picked data elements and their percent matches. The percent matches of compiler 2 ranged between ∼21% - 64% with compiler 1. This underscores the importance of data collected from multiple, trusted and standardized sources.

**Table 2:** Percent matches between overlapping elements of the two compilers.

| Variable name |  | No of categories | Percent match |
| --- | --- | --- | --- |
| Compiler 1 | Compiler 2 |  |  |
| Home Market Value Estimated | Home Value Range | 6 | 55.68% |
| Marital Status | Marital Status | 2 | 64.26% |
| Presence of Children | Presence of Children | 2 | 41% |
| Income Estimate Household | Income Range | 9 | 20.85% |

### 3.3 Bias

Underserved rural populations are less likely to get needed healthcare due to distance, costs, and poor insurance coverage leading to underdiagnosis of illnesses even though they are more commonly affected by chronic conditions as shown in **Table 3**. To study this problem of bias in healthcare data, we have analyzed and preprocessed a real-world data set of patients with chronic conditions from geographically disparate locations. We have chosen to preprocess because it prepares the data set for any following modeling and does not need to be repeated for each new classifier. We also tested the removal of the sensitive attribute altogether. Each of these tests produces similar AUC results. in **Table 3** or similar. (Seker, Talburt, and Greer 2022)

**Table 3:** As age increases, multimorbidity increases. However, in the case of rural residents a decrease in the average number of chronic conditions reflects bias which could impact patient care where predictive algorithms use this information.

|  | Geographic Location |  |
| --- | --- | --- |
| Age Groups | Rural | Urban |
| 18-64 | 2.3127 | 2.7463 |
| 65+ | 2.2340 | 3.0272 |

### 3.4 Preliminary Analysis and Visualization of the Merged Data

The SDOH data merged with EHR provided insights into the social risk factors of disease both at patient level as well as the population level. We first explored the disparities in demographics and SDOH factors between the 55,422 patients who were Covid positive compared to those who were not. **Table 4** provides a summary of a subset of patient characteristics that were compared between these two groups. Our preliminary analysis showed that apart from demographic factors, several SDOH factors like home-ownership, marital-status, presence of children, number of members per household, economic stability index and education were significant different between the two patient groups while estimated family-income and home market-value were not. **Figure 2** shows a map of the top COVID-affected zip codes in Arkansas overlaid on a heatmap image of average economic stability index of those zip codes. The darker blue counties were less economically stable and also had higher percentages of covid positive patients.

**Table 4:** Summary statistics of demographics and individual-level SDOH factors in the merged data.

| Characteristics | N | COVID Positive |  | p-val<sup>2</sup> |
| --- | --- | --- | --- | --- |
|  |  | No, N = 54330<sup>1</sup> | Yes, N = 1092<sup>1</sup> |  |
| **Gender** | 55422 |  |  | 0.032 |
| F |  | 29611 (55%) | 638 (58%) |  |
| M |  | 24706 (45%) | 454 (42%) |  |
| U |  | 13 (<0.1%) | 0 (0%) |  |
| **Age** | 55422 | 62 (18) | 46 (18) | <0.001 |
| **Race** | 46869 |  |  | <0.001 |
| African American |  | 10220 (22%) | 298 (32%) |  |
| Asian |  | 551 (1.2%) | 25 (2.7%) |  |
| Hispanic |  | 1445 (3.1%) | 110 (12%) |  |
| White/Other |  | 33726 (73%) | 494 (53%) |  |
| **Home Owner Renter** | 54068 |  |  | <0.001 |
| Home Owner |  | 34558 (65%) | 605 (56%) |  |
| Renter |  | 18420 (35%) | 485 (44%) |  |
| **Marital Status** | 54068 |  |  | <0.001 |
| Married |  | 26140 (49%) | 391 (36%) |  |
| Single |  | 26838 (51%) | 699 (64%) |  |
| **Has Children** | 54068 | 14826 (28%) | 377 (35%) | <0.001 |
| **Member Household** | 54068 |  |  | <0.001 |
| 1 |  | 20473 (39%) | 487 (45%) |  |
| 2 |  | 15603 (29%) | 224 (21%) |  |
| 3 |  | 7772 (15%) | 173 (16%) |  |
| 4 |  | 5201 (9.8%) | 112 (10%) |  |
| 5 |  | 3305 (6.2%) | 71 (6.5%) |  |
| 6 |  | 393 (0.7%) | 13 (1.2%) |  |
| 7 |  | 145 (0.3%) | 3 (0.3%) |  |
| 8 |  | 61 (0.1%) | 6 (0.6%) |  |
| 8Plus |  | 25 (<0.1%) | 1 (<0.1%) |  |
| **Estimated Family Income** | 34792 |  |  | 0.4 |
| Less30K |  | 0 (0%) | 0 (0%) |  |
| \$30-50K | | 14070 (41%) | 271 (43%) | |
| \$50-75K | | 9228 (27%) | 165 (26%) | |
| \$75-100K | | 4448 (13%) | 90 (14%) | |
| \$100-125K | | 2567 (7.5%) | 36 (5.7%) | |
| \$125Kplus | | 3848 (11%) | 69 (11%) | |
| **Estimated Home Market Value** | 51438 |  |  | 0.9 |
| Less100K |  | 19231 (38%) | 405 (39%) |  |
| 100-200K |  | 21183 (42%) | 434 (42%) |  |
| 200-300K |  | 5719 (11%) | 124 (12%) |  |
| 300-400K |  | 2153 (4.3%) | 39 (3.7%) |  |
| 400-500K |  | 878 (1.7%) | 20 (1.9%) |  |
| 500Kplus |  | 1230 (2.4%) | 22 (2.1%) |  |
| **Economic Stability Index** | 37135 |  |  | <0.001 |
| 10-15 |  | 7126 (20%) | 117 (15%) |  |
| 16-20 |  | 7696 (21%) | 118 (15%) |  |
| 21-25 |  | 10940 (30%) | 226 (28%) |  |
| 26-30 |  | 10573 (29%) | 339 (42%) |  |
| **Education** | 34295 |  |  | <0.001 |
| Attended Vocational/Technical |  | 361 (1.1%) | 5 (0.7%) |  |
| Completed College |  | 11392 (34%) | 187 (28%) |  |
| Completed Graduate School |  | 4008 (12%) | 45 (6.6%) |  |
| Completed High School |  | 17857 (53%) | 440 (65%) |  |

<sup>1</sup>Statistics presented: n (%); mean (SD)
<sup>2</sup>Statistical tests performed: chi-square test of independence; Wilcoxon rank-sum test

**Figure 2:**
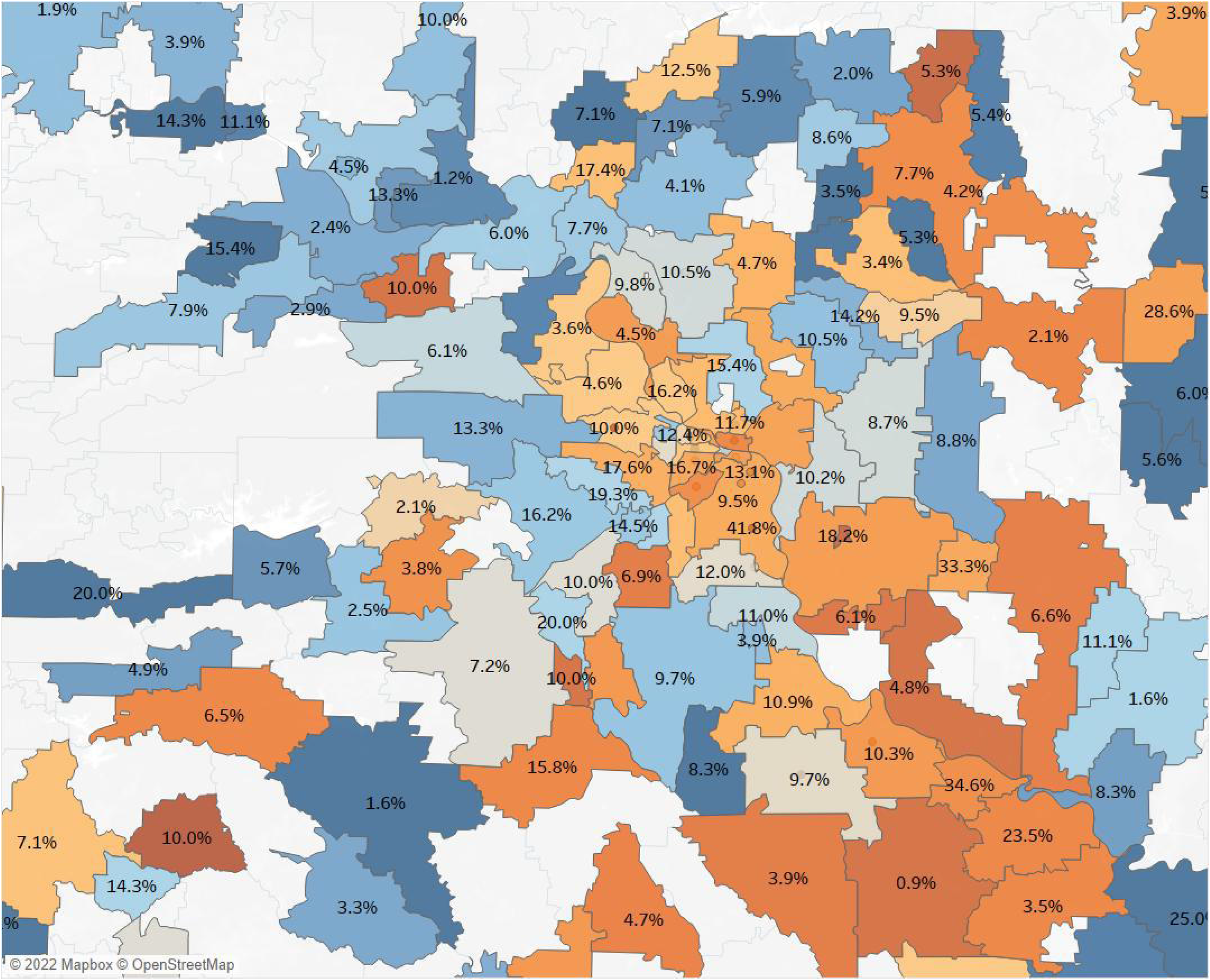
This map of central Arkansas shows that lower economic stability (indicated in orange) trends with higher instances of COVID-19 positivity (indicated in percentage label) but that was not true for all zip codes as there were other SDOH confounders.

## 4 Discussion

In this proof-of-concept study our main objective was to evaluate the viability of consumer marketing data, purchased from 3^rd^ party HIPPA-compliant vendors, as a source of SDOH factors that are largely missing from the EHR data. We purchased in-depth patient-level consumer data from two different vendors, mapped a wide array of data elements to the 5 broad SDOH domains as defined by the Healthy People 2030 SDOH framework, linked the consumer data to EHR patient level data, stored the data in a SQL server linked with several statistical and data exploration tools, evaluated data quality and preprocessed the data, and lastly completed a preliminary analysis of a subset of the SDOH elements that characterized the patients. To our knowledge this is the first study to explore the viability of consumer marketing data as a source of patient-level SDOH data.

With recent upsurge in research solidifying the significant relationship between SDOH and population health, an increasing number of healthcare stakeholders are exploring the use of public databases for community-level information in order to identify those patients that are most vulnerable to SDOH. For example, census tracts data have been used to identify areas associated with socio-economic risks and poor health outcomes.(Liaw et al. 2018) But a recent study by Cottrell et al (Cottrell et al. 2020) showed that only about 48% of the times community-level data can accurately identify social risks at the patient level. Thus, healthcare decisions on individual patients based on community-level data may fall short on providing adequate care to a significant number of patients. This may give rise to the problem of ‘ecologic fallacy’ where incorrect assumptions can be made about a patient based on aggregate-level information from community-level data. In this study we have attempted to address this problem by partnering with companies/vendors that are honed consumer market researchers.

We have developed a repeatable process to incorporate commercially compiled data into EHR data. The added value has been demonstrated based on a published paper (Greer et al. 2021) and other in-progress research efforts focused on disease specific predictive analytics. We have also identified opportunities in data quality research areas that need further study as part of this work. The curated data are being used to support several healthcare analytics applications, including descriptive analytics, data visualization, and predictive modeling. During this work, we have developed the first mapping scheme of commercial data elements with SDOH elements. This is a fascinating aspect of this work because the healthcare community has not reached a consensus on a standard set of social determinants of health concepts demanding that this process be agile and flexible. As we advance this area, our goal is to work on mapping and semantics using an ontology

Building an enrichment process for EHR data has allowed us to study important questions about temporal alignment, data dictionary and coverage issues, legal requirements, and security requirements. Initially, the legal, research and business processes required were complex and time-consuming. Patient data must be kept private and secure at all times, and all parties must be bound by a contract to minimize the possibility of a data breach. Fortunately, once contracts and transfer processes are in place, they remain active and available for repeated consumer data collection. This is important because continued collection will be necessary. Compiled data becomes stale over time, and EHR data is collected only at the time of each encounter. Aligning these time windows is necessary for elements that must be current while is less critical for elements that are more likely to remain stable over time. As the process iterates and newly compiled data is integrated into the EHR data, the data dictionary must also be updated. We discovered that the data dictionaries provided by compilers vary in quality and detail. In addition, compilers are continuously adding, removing, and updating elements resulting in multiple versions of the dictionary documentation. Integrating data from these dynamic systems also impacted the mapping of compiled elements onto SDOH concepts, resulting in a mapping component for each iteration. If kept current, the SDOH mapping will require minimal effort to maintain. Throughout all of these components, it was also necessary to tackle practical Information technology issues such as storage, tools, permissions, and access which will need to be customized to each institution.

Our study had several limitations. Due to budgetary constraints we restricted our SDOH data sources to two different vendors only. One had more mapped data elements while the other had more coverage, thus highlighting the need for data collection from multiple, trust worthy and reputable vendors with standardized methods of data collection. There were significant missing data in each data elements. Also, among the overlapping data elements, the concordance between the data was not very high which underscores the need for good quality data sources. May be a standardized and validated screening tool based on our approach needs to be developed that can be used by all healthcare entities.

In conclusion, we have developed a repeatable SDoH enhancement process to incorporate dynamically evolving SDOH domain concepts from consumers into clinical data. The literature provides early and rapidly growing evidence that integrating individual-level SDoH into EHRs can assist in risk assessment and predicting healthcare utilization and health outcomes, which further motivates efforts to collect and standardize patient-level SDoH information. This study highlights one potential means to incorporate individual-level patient data into EHR, thus opening up possibilities for predictive analytics and enhanced solutions for providers, payers and healthcare organizations to enable them to address the social needs of patients.

## Data Availability

No data produced in the present study are available

## 6 Author Contributions

MG developed the concept. MG SB worked on concept development analysis and manuscript writing. CZ performed SDOH mapping and writing.

